# Active Engagement: The Impact of Group-Based Physical Activities on Resilience of Israeli Adolescents with ADHD

**DOI:** 10.1101/2024.01.09.24301060

**Authors:** Yair Tamir, Anne Marie Novak, Itzhak Cohen, Bruria Adini, Shahar Lev-Ari

## Abstract

**Background:** Attention Deficit Hyperactivity Disorder (ADHD) is a pressing concern in contemporary pediatric public health, with its prevalence rising among children and teenagers. This study explored the relationship between group-based physical activity and the well-being, resilience, and distress levels of Israeli youth, with a specific focus on those with ADHD.

**Methods:** This was a survey-based, cross-sectional study that included 699 Jewish Israeli teenagers aged 16 to 19. In addition to quantitative questionnaires that examined sociodemographic factors, resilience, distress, and well-being levels, the youth were asked about participation in group-based physical activities and the significance they ascribe to various facets of the activities.

**Results:** Our findings indicated that structured and group-based physical activities, especially the Five Fingers program which emphasizes psychosocial development and leadership skills, are associated with higher resilience (p<.01) and lower distress levels (p<.01) in adolescents. Generally, Israeli adolescents with ADHD exhibited lower levels of resilience (p<.001) and well-being (p<.001), and higher levels of distress (p<.001) compared to their counterparts. However, adolescents with ADHD who participated in group-based activities fared better in terms of distress (p<.01) and well-being (p=.018) than adolescents with ADHD who did not participate in organized sports. Further, participation in any form of sport activity, older age, male gender, and a higher socio-economic status predicted greater resilience in youth generally.

**Conclusions:** The study presents the potential of structured, engaging physical activities that involve psychosocial training and group integration activities to improve the mental health of adolescents, especially in the context of ADHD.

## Introduction

Attention Deficit Hyperactivity Disorder (ADHD) remains one of the most urgent concerns for contemporary pediatric public health, with the most recent meta-analysis estimating its prevalence in children at 7.6% and 5.6% in teenagers [1]. The incidence of ADHD is on the rise [2], at least in part due to the increased awareness and reduced stigma among physicians and the general public, leading to an increase in reported diagnoses [3]. ADHD can significantly impact academic performance [4], social competence [5], and overall well-being [6] during a crucial developmental phase. Currently, various strategies are employed to address ADHD in adolescents, including medication, behavioral therapy, and educational support [7]. However, significant gaps remain in our understanding of the disorder and its optimal management. In recent years, emerging research suggests that physical activity could play a pivotal role in mitigating the symptoms of ADHD in teenagers [8] offering a promising avenue for further exploration and potential intervention. By delving deeper into the relationship between exercise and ADHD, we may uncover novel and effective ways to enhance the lives of adolescents grappling with this neurodevelopmental disorder.

Research suggests that physical activity is beneficial for overall ADHD symptoms, executive function and motor abilities, with possible additional benefits to one’s social, emotional, and behavioral health [9]. It has been hypothesized that physical activity may have an effect on neurodevelopment, through its effect on neurotransmitters and neurogenesis, as well as enhanced blood flow to the brain and growth in brain tissue volume [10,11]. Very few studies examined the relationships between physical activity and the well-being, resilience, and distress levels of people with ADHD, and adolescents in particular. This gap in knowledge must be addressed: adolescents with ADHD have been shown to suffer from increased levels of stress [12] as well as reduced well-being and resilience [6]. A 2016 review on the effect of physical activity on children with ADHD identified five moderators between the activity and the outcomes: the type of the activity, its intensity and length, its duration, and its frequency [8]. Furthermore, the structurization of the physical activity, and performing it in an integrated and cohesive group may play a role in the ability of the individual to consistently engage in it [13,14].

Established in Israel in 2014, *Five Fingers* (*Khamesh Etzba’ot*) is a unique educational and physical activity organization with a strong focus on fostering cultural and social impacts on lifestyle habits, resilience, self-perception, meaning, belonging, and leadership among adolescents [15]. The program emphasizes psychosocial development through physically challenging activities, team-building and social cohesion exercises (or physical-mental group training, as defined by the organization). Its primary goals include self-development, building self-trust and trust in others, and cultivating leadership skills, both in times of adversity and success. Within the Five Fingers program, factors such as debriefing, team camaraderie, coach/instructor relationships, and significant organizational events take on distinctive importance. The culture of debriefing encourages a pragmatic approach to both failures and successes, fostering personal growth and achievement. Israeli adolescents who participated in online *Five Fingers* physical-mental group training during the COVID-19-related lockdowns, had higher resilience, satisfaction with life and better coping skills than those who did not, and the results suggested the importance of the structure and nature of the activity for the mental health of the participants [15]. To the best of our knowledge, this type of engagement in physical activity in adolescents with ADHD and its implications have not yet been addressed in research.

This study aimed to assess the resilience, well-being and distress levels of the Israeli youth, and identify the variables that predict their resilience, in particular in Israeli youth with ADHD. Furthermore, we aimed to measure the relationship between these factors and participation in different types of sport activities: unorganized, structured, and physical-mental group training (Five Fingers). We hypothesized that age, gender, and one’s socio-economic status as well as the presence of ADHD would be related to the degree of resilience. Furthermore, we hypothesized that the type of sport activity one is engaged in would be related to the investigated variables, with the subjects who participate in physical-mental group training experiencing the lowest levels of distress and highest levels of resilience and well-being.

## Methods

### Study design

This was a survey-based, cross-sectional study conducted through an Internet platform.

### Study setting and population

The study included 699 Jewish Israeli teenagers aged 16 to 19 living in central and northern Israel, who had either completed high school education or were high school students at the time, prior to the mandatory conscription into the Israeli army. The subjects were recruited by iPanel, the largest online panel operating in Israel, and 196 subjects who participated in the *Five Fingers* activities were recruited from their network through its channels. All subjects filled out an informed consent form to participate in the study or, if underage, presented a form signed by their guardian. The subjects were not aware of the central theme being investigated, to prevent possible bias in the subjects’ answers. The recruitment took place between the 10^th^ and the 20^th^ of May, 2021. The participants filled out an online questionnaire through the online platform.

### Study Tools and Outcome Measures

The independent variables in the study included gender, age, the socio-economic status, district of residence, religiosity, presence of ADHD symptoms and time devoted to a sport activity. Age was divided into three categories: 16, 17, or 18-19. The participants were asked to define their socio-economic status as either below-average, average, and above-average. The place of residence area was defined as either Haifa and northern Israel, Tel Aviv and Gush Dan (the central district), or the Sharon district. The level of religiosity was measured on a four-level scale: secular, traditional, religious, and ultra-Orthodox.

The presence of ADHD symptoms was identified using the 6-item (Part A) World Health Organization Adult ADHD Self-Report Scale (ASRS), which has been in wide use since 2005 [16]. The scale has high internal consistency and validity, and the possible scores range from 0 to 6, with scores of 4 or more indicating consistency with an ADHD diagnosis in adults [16]. Clinicians and researchers utilize the scale for older adolescents and adults alike [17]. As such, a score of 4 or more was considered indicative of ADHD for the purpose of the current study. In the current study, the ASRS reached an Alpha Cronbach score of 0.72, indicating acceptable internal consistency.

The main independent variable in the study, participation in a physical activity group, was self-assessed as either “not participating in any group physical activities,” “participating in unorganized sport activities,” “participating in a structured group physical activity,” and “participating in Five Fingers activities.” The participants were asked to grade the level of significance that they ascribe to various aspects of the activity group that they partake in content of physical activity, session initiation and closure discussions, intra-group friendships, relationship with the coach, and the special events of the program. The variable values ranged from 1 (not important at all) to 5 (very important). This questionnaire was developed and validated in 2020 for a study on adolescent participation in sports programs at the peak of the COVID-19 pandemic in Israel [15], and had acceptable internal consistency in the current study (Cronbach α score of 0.77).

The dependent variables of the study included distress, resilience, and well-being. The sub-scales of anxiety and depression, derived from the Brief Symptom Inventory (BSI) were employed for the measurement of distress (BSI) [18,19]. The five items of the depression subscale pertain to a bad mood, loneliness, lack of interest in daily activities, feelings of worthlessness, and hopelessness. The four items anxiety subscale refer to perceived nervousness, tension, and restlessness. Each item was rated on a scale ranging from 1 (not suffering at all) to 5 (suffering very much). Reliability for both subscales were high (α = 0.90). The overall questionnaire had good internal consistency (Cronbach α of 0.86).

The Connor–Davidson resilience scale (CD-RISC-10), which has been previously validated for this population [15], was used to assess resilience. The ten-items are phrased so that a higher endorsement of a statement indicates higher resilience (0 = not at all true, 1 = rarely true, 2 = sometimes true, 3 = often true, and 4 = true nearly all the time). People with lower-resilience categories tend to rate individual items in the 0–2 range, those with medium resilience tend to rate items as 3, and those with higher resilience tend to rate items as 4. A total of ten items were presented; CD-RISC scores range from 0 to 40. The questionnaire had good internal consistency (Cronbach α of 0.87). Well-being was quantified using a nine-step questionnaire that assesses the individuals’ perceptions of their lives in various contexts, including health, free time, family, and others as previously described [20]. The subjects were asked to rate their degree of agreement with each comment on a scale of 1 (not at all) to 5 (very strongly). The questionnaire was found to be reliable (Cronbach α = 0.84). The study was approved by the Tel Aviv University Review Board (Approval 0002997-2, 4/2021).

### Statistical Analysis

All study data were analyzed using SPSS software, version 27.0. Statistical significance was defined as p≤0.05. The minimum sample size for the required statistical tests was calculated using G*Power3 [21], to test for the differences in distress, well-being and resilience of different sport activity groups, and stood at 300 participants. As such, given the large study population (N=699), the statistical power for this study stood at 0.99.

We used descriptive statistics to assess the distribution of the socio-demographic characteristics of the subjects. The data were assessed using Chi-square tests and Cramér’s V values. To examine the difference in distress, well-being, and resilience of the subjects in the physical activity groups, three one-way Anova tests were conducted. Effect sizes were calculated to assess the effect of the trial groups on distress, well-being, and resilience averages. To examine the origin of the mean difference between the trial groups, follow-up tests with Bonferonni correction were conducted between each of the two groups.

For subjects in test groups 2 and 3 of which members were engaged in some kind of social activity, independent t-tests were conducted to examine the difference in the averages of each group trait. The effect sizes for the test group’s effect on the variable values were calculated using Cohen’s effect size. In addition, correlations between the values of the scales were calculated, without division into the experimental groups.

Finally, linear regression models were created to examine the combined effect of the study variables on predicting the extent of the subjects’ resilience. In order to estimate the pure effect of each variable on participant’s resilience, we used a hierarchical model, which considers all the variables that were found to be significantly different while using the univariate tests. This method calculates the effect of each independent variable on resilience, standardized to all other variables that are shown in the model. The regressions were calculated separately for subjects who did not engage in any group physical activity and separately for subjects in the trial groups 2 and 3 who engaged in group physical activity. For each group, the first regression model presented examines the effect of all variables on the resilience of subjects and the second model includes the minimum number of predictors required to predict the degree of resilience of subjects. In total, four multivariable regression models are presented in this paper.

## Results

### The Characteristics of the Study Group

The study included 699 Israeli teenagers aged 16 to 19, before the conscription into mandatory military service. Less than a quarter of the subjects (21.3%) were aged 16, less than a third were aged 17 (29.3%), and about half of the subjects were 18 or older (49.4%). Most of the subjects in the study were secular (63%) and came from higher-than-average socioeconomic backgrounds (55%). About 17% of the subjects reported experiencing at least four ADHD symptoms.

Table 1 displays the distribution of the independent variables of the study: gender, age, socio-economic status, presence of ADHD, time devoted to a sports activity, location, and religiousness in relation to the three sports activity groups that were examined.

**Table 1.**
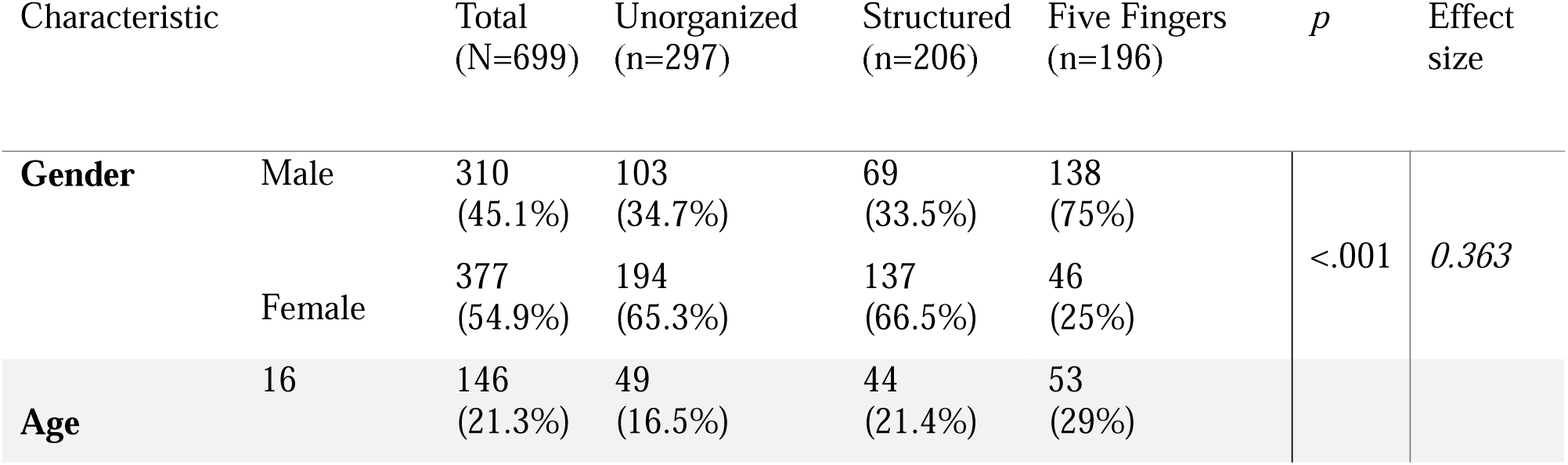
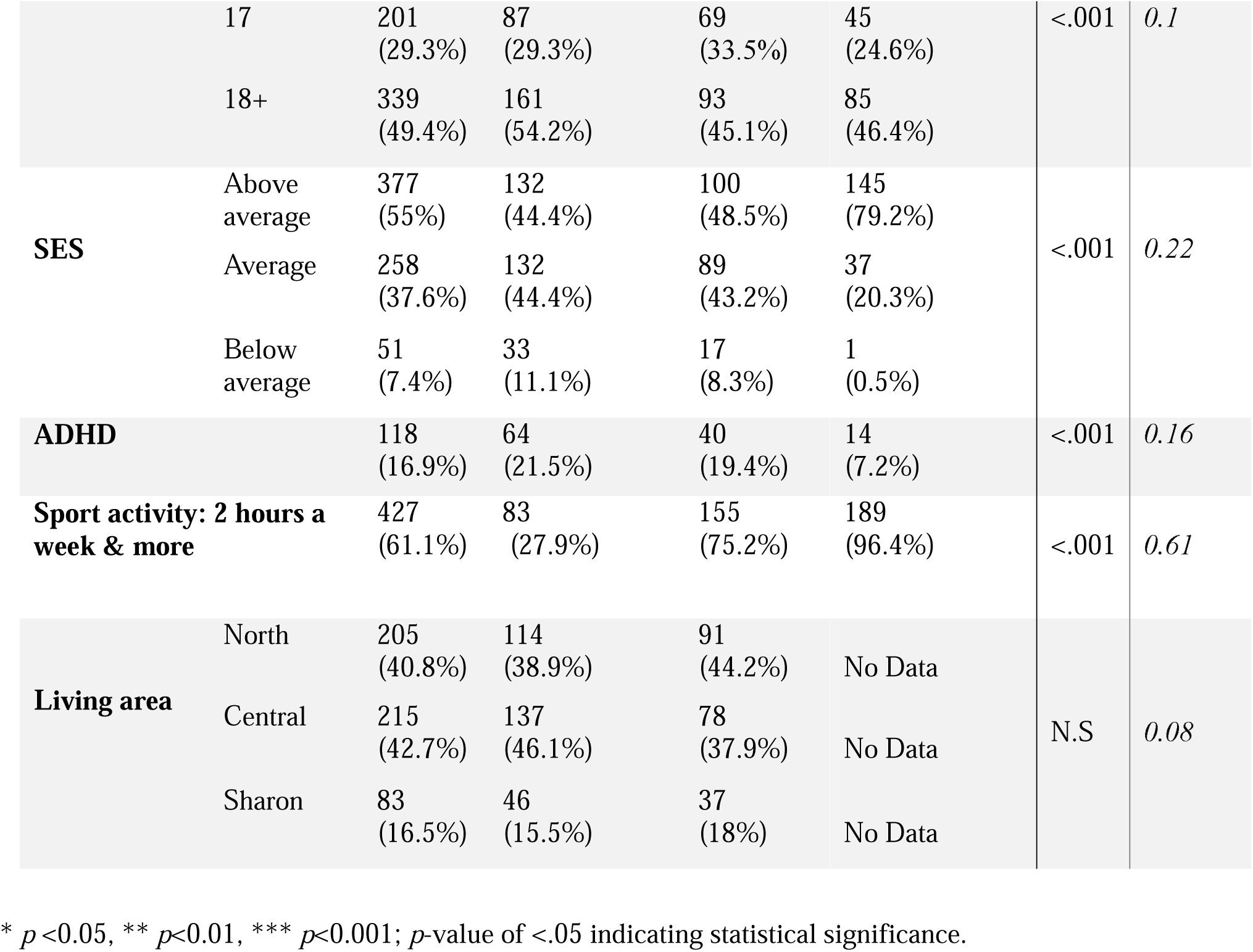
Study population.

### Linear Regression Models for the prediction of Resilience

The multi-variable regression models included the variables that may affect the subjects’ resilience. Table 2 presents the initial regression models that included solely the background characteristics (see Table 2) of those participants who did not engage in group physical activity. Table 3 presents additional two models that included several variables that could partially explain the subjects’ resilience.

**Table 2.**
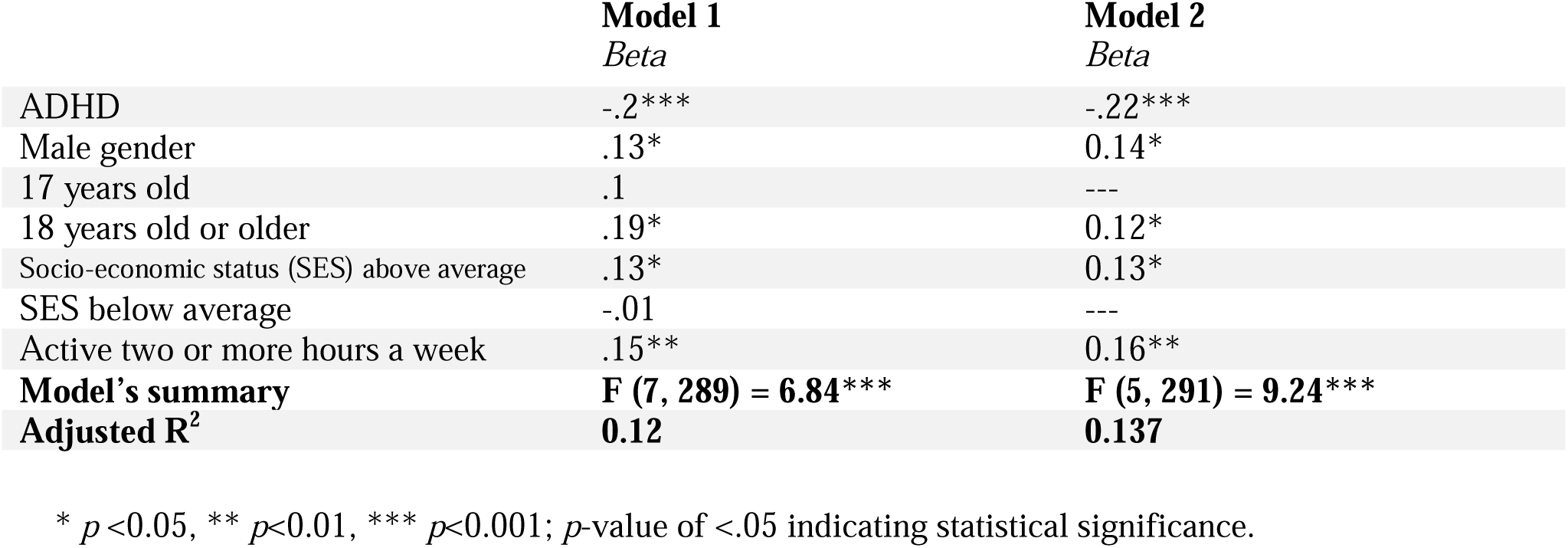
Linear Regression Models for the prediction of Resilience in Participants who did not engage in group-based physical activity: Background Characteristics.

|  | **Model 1**<br>*Beta* | **Model 2**<br>*Beta* |
| --- | --- | --- |
| ADHD | -.2*** | -.22*** |
| Male gender | .13* | 0.14* |
| 17 years old | .1 | --- |
| 18 years old or older | .19* | 0.12* |
| Socio-economic status (SES) above average | .13* | 0.13* |
| SES below average | -.01 | --- |
| Active two or more hours a week | .15** | 0.16** |
| **Model's summary** | **F (7, 289) = 6.84\*\*\*** | **F (5, 291) = 9.24\*\*\*** |
| **Adjusted R<sup>2</sup>** | **0.12** | **0.137** |

\* $p < 0.05$ , \*\* $p < 0.01$ , \*\*\* $p < 0.001$ ; $p$ -value of $< .05$ indicating statistical significance.

**Table 3.** Linear Regression Models for the prediction of Resilience in Participants who engage in physical activity: all explanatory variables.

|  | **Model 1**<br>*Beta* | **Model 2**<br>*Beta* |
| --- | --- | --- |
| Physical Activity Group (Five Fingers) | .15** | .18*** |
| ADHD | -.17*** | -.16*** |
| Male gender | .05 | ----- |
| 17 years of age | .16** | .16** |
| 18 years of age or older | .2*** | .21*** |
| Socio-economic status (SES) above average | .16*** | .16*** |
| SES below average | .004 | ----- |
| Active two or more hours a week | -.07 | ----- |
| Content of physical exercise | .19*** | .19*** |
| Initiation and closure discussions | .06 | ----- |
| Intra-group friendships | .04 | ----- |
| Relationship with coach | .6** | .18*** |
| Program special events | -.03 | ----- |
| **Model's summary** | **F (13, 376) = 12.18\*\*\*** | **F (7, 382) = 21.98\*\*\*** |
| **Adjusted R<sup>2</sup>** | **0.26** | **0.28** |

\* $p < 0.05$ , \*\* $p < 0.01$ , \*\*\* $p < 0.001$ ; $p$ -value of $< .05$ indicating statistical significance.

All socio-demographic variables which were found significant in Table 1 are included in Model 1 Regressions, followed by Model 2 Regressions which include only variables which were found to have a significant relationship with the participant’s resilience.

The degree of resilience of youth aged 17 was not significantly different from younger teenagers, who represented the reference group for the calculation presented in Table 2. Additionally, the degree of resilience of subjects from lower-than-average socioeconomic backgrounds was not significantly different from those of an average socioeconomic status. ADHD was found to have the greatest impact on the subjects’ resilience: subjects with four or more ADHD symptoms had lower resilience compared to the other subjects (p<0.001). Weekly exercise of at least two hours per week was found to have a positive effect on the level of resilience of the subjects (p<0.01).

Boys were found to have higher resilience than girls (p<0.05). Subjects from higher-than-average socioeconomic backgrounds were found to have higher resilience compared to those from average socioeconomic backgrounds (p<0.05). In addition, a significant positive effect of age was found on the level of resilience of the subjects, so that subjects aged 18 and older were found to have higher resilience compared to subjects aged 16. A model that uses these five variables to predict the level of resilience of the subjects explained about 14% of the variance in resilience (p<.001).

We built different models to predict the degree of resilience of the subjects that engaged in a structured group physical activity and for those engaging in Five Fingers group activities. For the latter, additional data was gathered regarding the importance they attached to various aspects of the group activity. The findings are presented in Table 3.

Among subjects who participated in group-based physical activities, participants who were members of the "Five Finger" groups were found to have higher level of resilience, compared to participants practicing in other sport activities (*p*<.001). The level of resilience among youth aged 17 and 18 and older was higher than among subjects aged 16 (*p*<.01). In contrast to those who did not engage in group exercise, we found that one’s gender and the length of activity was not related to one’s resilience. Similarly to the findings in the non-exercising group, the socioeconomic status was found to have a positive effect on the level of resilience of the subjects, with subjects from higher-than-average socioeconomic backgrounds having higher resilience compared to subjects from average socioeconomic backgrounds (p<.001). In addition, ADHD had a negative effect on the subjects’ resilience (p<.001). Of the criteria tested, the content of exercise (p<.001) and the relationship with the coach (p<.01) appeared to be related to the resilience of subjects who engaged in group-based physical activity. A smaller model, comprising only the statistically significant variables, included age, SES, ADHD, training group style, relationship with the coach, and the content of physical exercise, and explained approximately 28% of the variance of the resilience.

An additional interaction model was constructed to examine the interaction between practicing in "Five Fingers" group and ADHD symptoms on one’s resilience. The interaction variable (*Beta* = 0.1, *p*<.05) was additional to all seven variables included in Model 2 regression. The model was found significant: F (8, 381) = 19.83, p<.001, with adjusted R^2^ of 0.29. This finding suggests the effect of practicing in "Five Fingers" group on one’s resilience is bigger among youth with ADHD (*p*<.05). In other words, while the effect of practicing in "Five Fingers" group on non-ADHD youth is 0.18, the effect of practicing in "Five Fingers" group on ADHD youth is 0.28 (p<.05).

Testing the study’s hypotheses on differences in distress averages, resilience and well-being was performed using three tests to analyze one-way variability for each of the dependent variables. We found a distinct difference in the distress, resilience, and well-being values of the subjects in the groups, and thus post-hoc tests were conducted to detect the differences between the different training groups. The findings of the follow-up analyses are presented in Figure 1. Post-hoc tests with Bonferonni corrections to examine the source of the difference between the groups indicate that the group participating in Five Fingers activities experienced the lowest distress levels among the three test groups (p<0.01). The subjects in this group were also found to have the highest resilience and levels of well-being as compared to the rest of the subjects (p<0.01).

**Figure 1.**
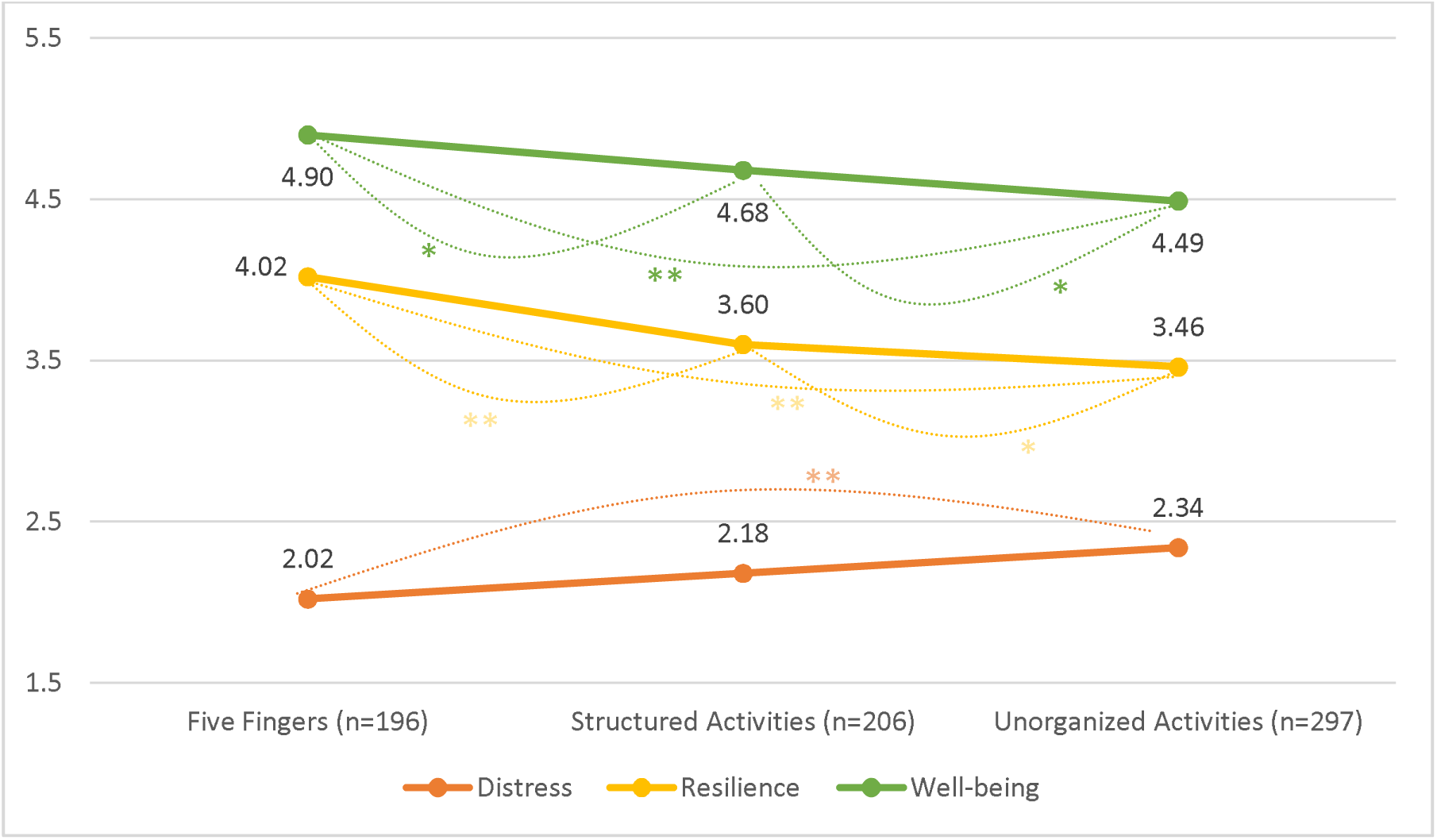
Difference in distress, resilience, and well-being in the study groups. The differences in the examined variables: distress (in orange), resilience (in yellow), and well-being (in green) between the three types of sport activities (Five Fingers, structured, unorganized). Statistical significance denoted as p-value less than than 0.01 (**) or less than 0.05 (*). The displayed values are the mean scores.

Figure 2 shows that in all five criteria for the evaluation of the meaningfulness of different aspects of the activities, the Five Fingers group subjects indicated higher values compared to the participants in other structured group activities. All of the differences between these two groups were significant (p<0.001).

**Figure 2.**
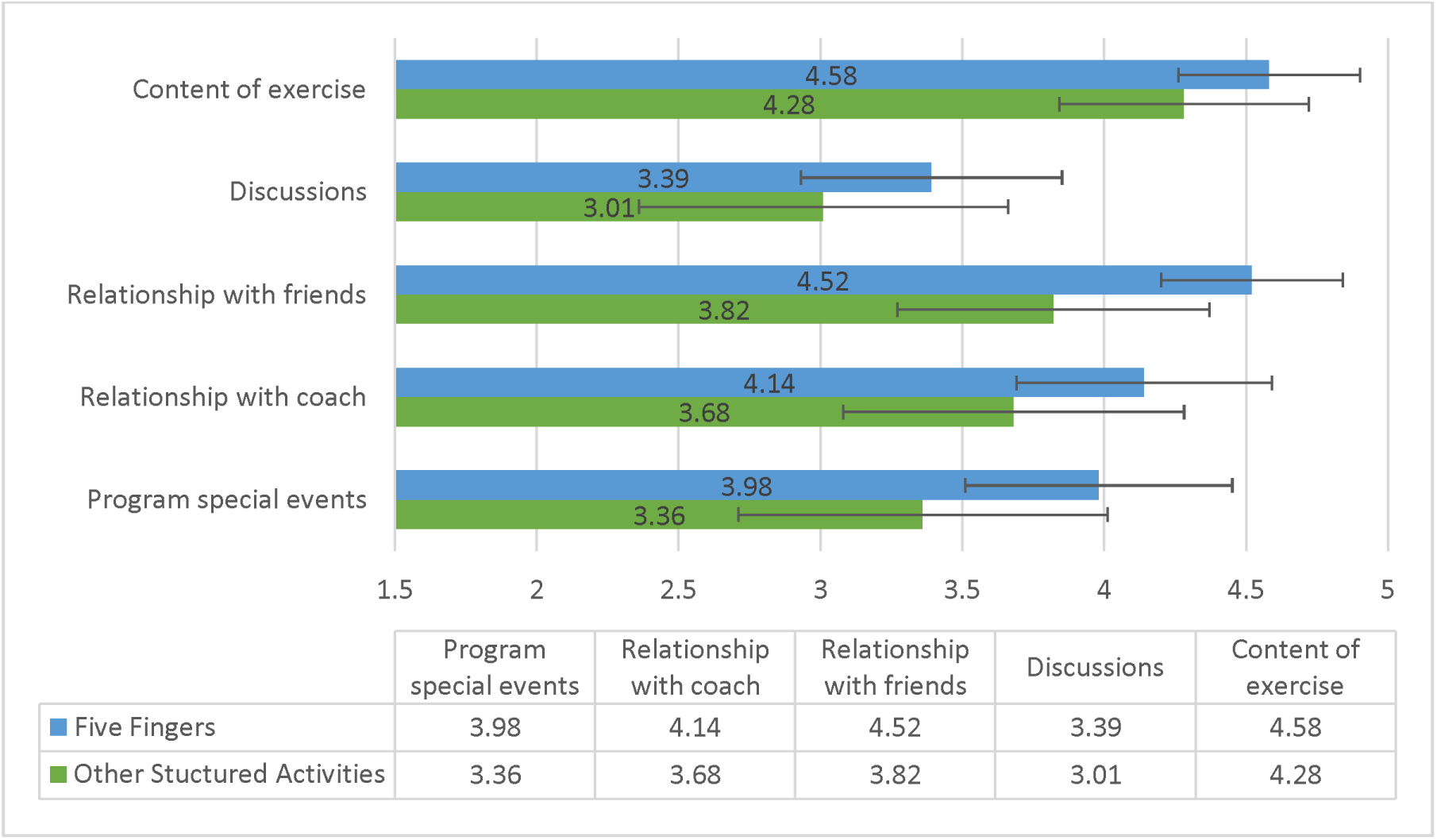
Difference in the meaningfulness of activities as assessed by participants in group physical activities.

Table 4 presents the correlations between the factors examined in the participants in group sport activities. The strongest correlations were observed between special events and coaching (.549, p<.01), friendships (.505, p<.01) and debriefing (.535, p<.01), as well as friendships and coaching (.536, p<.01). All of the correlations presented reached statistical significance (p<.01).

**Table 4.**
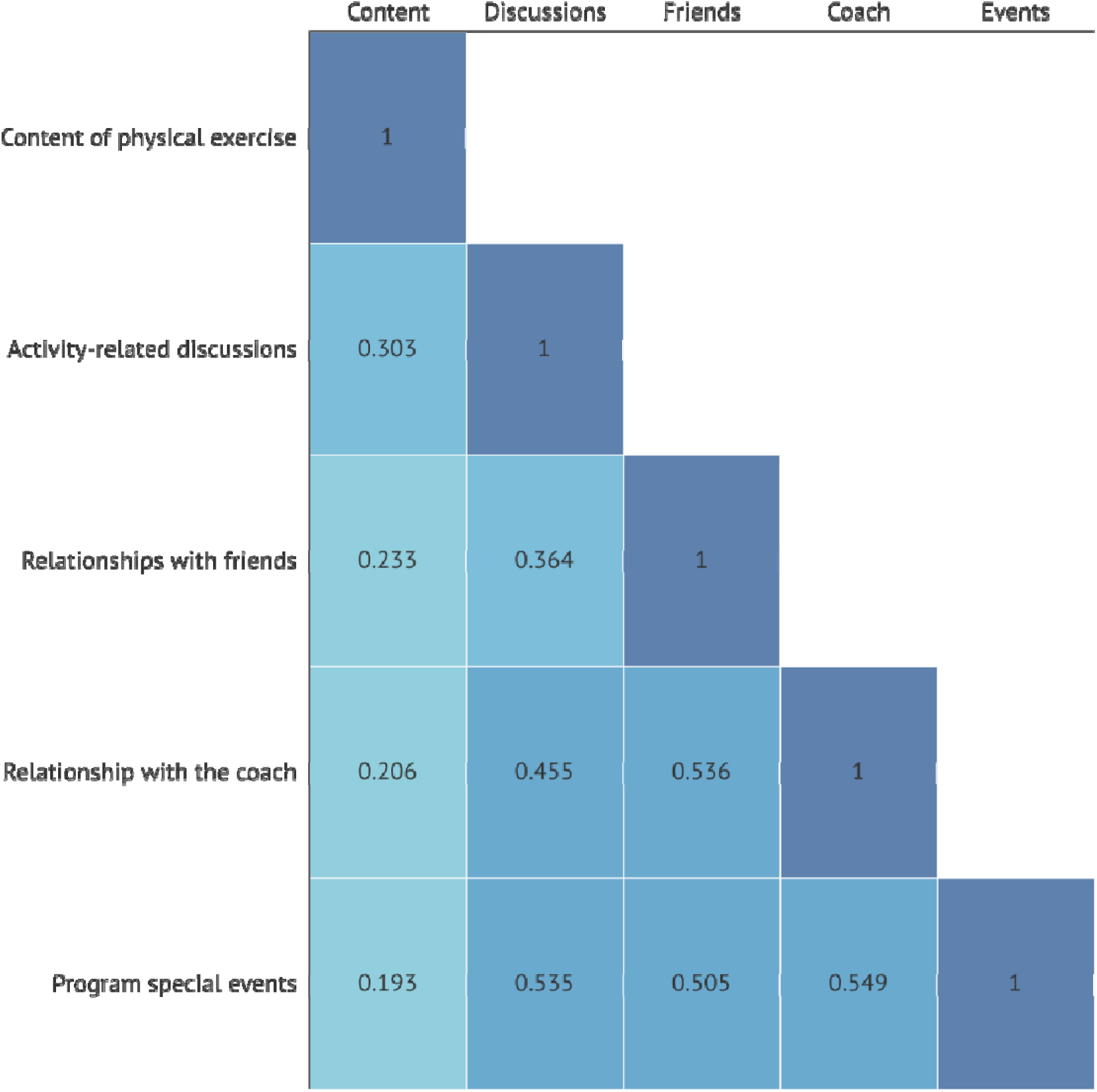
Correlations between the meaningfulness of activities in the group-based physical activities. The figure presents the correlations between the five characteristics of group sport activities that the participants may attach meaning to. All the correlations were statistically significant.

### Distress, Resilience, and Well-being among adolescents classified with ADHD

Table 5 presents the differences in the scores for distress, resilience, and well-being among adolescents classified as suffering from ADHD (ASRS score of 4 or higher) next to their counterparts. All study participants with ADHD had significantly lower resilience (p<.001, d=.78) and well-being (p<.001, d=.9) and higher levels of distress. (p<.001, d=1.05). In adolescents taking part in the Five Fingers activities, we observed significant differences in distress (p<.001) and well-being (p=.018) between ADHD and non-ADHD participants, but we observed no difference between the two groups in resilience. Adolescents with ADHD who participated in the Five Fingers activities lower values in the activity they engaged in (p<.001), debriefing (p<.001) and the program special events (p=.022) than their counterparts.

**Table 5.** Distress, Resilience, and Well-being in the Participants with and without ADHD.

| Factor |  | Total<br>(N=699) | Mean (SD) | *p* | *t* | Effect size (*d*) |
| --- | --- | --- | --- | --- | --- | --- |
| **Distress** | Non-ADHD | 581 | 2.07 (0.72) | <.001 | 10.35 | 1.05 |
|  | ADHD | 118 | 2.85 (0.84) |  |  |  |
| **Resilience** | Non-ADHD | 581 | 3.75 (0.64) | <.001 | 7.71 | 0.78 |
|  | ADHD | 118 | 3.23 (0.72) |  |  |  |
| **Well-being** | Non-ADHD | 581 | 4.78 (0.74) | <.001 | 8.96 | 0.9 |
|  | ADHD | 118 | 4.1 (0.8) |  |  |  |

Independent t-tests were conducted to examine whether mean differences in the significance attached by the participants to characteristics of the activities (see Table 4) are related to ADHD among the 390 participants who exercised regularly. Our results showed that participants with ADHD attached less importance to these characteristics, and had consistently lower means for each, compared to non-ADHD participants. Participants with ADHD were found to have significantly lower means in content of physical exercise [(3.96 vs. 4.49), t (388) = 4.74, p<.001], initiation and closure discussions [(2.75 vs. 3.57), t (388) = 4.5, p<.001] and program special events [(3.33 vs 3.71), t (388) = 2.08, p<.05].

## Discussion

Our study contributes to the expanding body of research on the associations between resilience, distress, well-being, and physical activity in the context of adolescent ADHD. Israeli adolescents with ADHD exhibited lower levels of resilience (p<.001, d=0.78) and well-being (p<.001, d=0.9), and higher levels of distress (p<.001, d=1.05) compared to their non-ADHD counterparts. Our analyses further revealed that physical activity was related to greater resilience among all study participants, which was further related to their gender (with boys having greater resilience than girls), socio-economic status, and age (with older participants exhibiting greater resilience). Our study distinguished between various types of physical activity and found that those engaging in structured activities, as opposed to unorganized ones, exhibited greater well-being (p<.01), greater resilience (p<.01), and lower distress levels (p<.01). Notably, the Five Fingers program, an Israeli educational and sports organization focusing on team building, self-development, and physical-mental training, yielded the most favorable outcomes. Furthermore, exercising with "Five Fingers" was found to increase resilience in the study participants with ADHD symptoms more than in those without (p<.05). Participants in the Five Fingers program attached more significance to different aspects of group physical activity, including exercise content, discussions, relationships with coaches and friends, and special events. Adolescents with ADHD who participated in the Five Fingers activities reported lower distress levels and higher well-being compared to their non-ADHD peers, although they attributed less significance to the various aspects of the program.

Our findings align with previous research demonstrating that physical activity can have a positive impact on the lives of teenagers with ADHD. Gapin et al. reported that exercise can enhance cognitive function and reduce ADHD symptoms in children and adolescents [22], while Hoza et al. conducted a meta-analysis showing that physical activity interventions can improve attention, hyperactivity, and impulsivity in children with ADHD [9]. Furthermore, it was shown that group-based physical activities specifically protect against depression [23]. This is consistent with earlier research demonstrating that social support and social connectedness can positively influence mental health outcomes [24,25]. Our study extends this literature by examining the role of group-based physical activity in the context of ADHD in teenagers in particular, revealing that such engagement was associated with higher levels of resilience (p<.001, d=0.78), well-being (p<.001, d=0.9), and lower distress (p<.001, d=1.05). Further, our analysis revealed that exercising with the Five Fingers group increased resilience of the youth with ADHD more so than in those without ADHD symptoms.

The study highlights a noteworthy association between religiousness and sports activity groups. Secular participants (62.8%) displayed higher involvement in sports activities compared to those with traditional (13.4%) and religious (6.6%) backgrounds. Religiousness is not a variable that has been widely examined in existing research on physical activity. However, it is possible that religiousness may be related to other factors that have been shown to impact mental health outcomes, such as social support and coping strategies [26]. It is plausible that religious commitments and traditions influence sports participation, including one’s availability during the weekend and the holidays, but the practical impacts of organized religion on youth sports activities remains largely unexplored, and merits further investigation.

The presence of ADHD symptoms is significantly related to sports activity groups, with a larger proportion of participants without ADHD (61.1%) engaging in sports activities compared to those with ADHD symptoms. Individuals with ADHD may face challenges in participating in organized sports and physical activities [8]: people affected by ADHD tend to experience greater difficulties in maintaining a schedule and staying committed [27]. Previous studies on this population showcased that attaching meaning and creating a sense of togetherness in activities enhances the engagement and performance of people with ADHD [28]. However, previous research has not explored how ADHD affects the perceived meaning or significance of sports activities. Our study specifically analyzed how a sport activity that devotes special attention to team-building and meaning-creation impacts adolescents, revealing that adolescents with ADHD who engaged in these activities reported lower levels of distress (p<.01) and greater well-being (p=.018). However, they attributed less meaning to debriefing, type of activity, and special events compared to their non-ADHD peers, suggesting distinct patterns of engagement. This aspect of the study results suggests that ADHD may have nuanced effects on the subjective experiences of adolescents during physical activities.

Several limitations of our study should be acknowledged. First, the cross-sectional design employed in this research, while providing valuable insights into the associations between resilience, well-being, distress, and physical activity in youth with ADHD, limits our ability to establish causality. Longitudinal studies would be necessary to further explore the directionality of these relationships. Nonetheless, it is important to note that we recruited a large sample of 699 participants and used regression analysis to predict the relationships between variables within the dataset to enhance the robustness of our findings. Second, the use of the Adult ADHD Self-Report Scale (ASRS) questionnaire for the identification of ADHD, although a well-validated and established tool, does not constitute the most comprehensive means of identifying ADHD. Clinical interviews and comprehensive assessments would provide a more definitive diagnosis of ADHD, and future studies may benefit from incorporating such measures. Lastly, sociodemographic differences between groups could impact the relationships observed in our study. To address these differences, we used a hierarchical model in our analysis, which considers all the variables that were found to be significantly different in the univariate tests.

## Conclusion

Attention Deficit Hyperactivity Disorder (ADHD) poses a significant challenge for contemporary pediatric public health, with its prevalence on the rise, particularly among teenagers. Despite the substantial impact of ADHD on various aspects of adolescents’ lives, including academic performance, social competence, and overall well-being, there remain gaps in our understanding of the disorder and effective management strategies. Recent research indicates that physical activity may contribute toward mitigating the symptoms of ADHD, with potential benefits for social, emotional, and behavioral health. While existing studies have primarily focused on the relationship between physical activity and executive function and motor abilities, this study delved deeper into the connections between exercise and well-being, resilience, and distress levels in adolescents, particularly those with ADHD. The findings revealed the significance of structured and group-oriented physical activities, such as the Five Fingers program, which places emphasis on fostering psychosocial development, self-development, and leadership skills. This unique approach to physical activity is associated with increased resilience and lower levels of distress, highlighting its potential for improving the mental health of participants. Our study aimed to identify key variables related to the resilience, well-being, and distress levels of Israeli youth, with a particular focus on those with ADHD, and to explore the association between these factors and participation in various types of sport activities. We observed that age, gender, socio-economic status, religiousness, and the presence of ADHD were all linked to varying degrees of resilience, distress, and well-being, while the type of sport activity engaged in played a role in determining these outcomes. Our findings underscore the potential of structured and socially engaging physical activities to positively impact the well-being of adolescents, shedding light on a promising avenue for addressing ADHD in this age group.

## Data Availability

All data produced in the present study are available upon reasonable request to the authors

